# Linking vestibular function and sub-cortical grey matter volume changes in a longitudinal study of aging adults

**DOI:** 10.1101/2020.11.17.20233684

**Authors:** Dominic Padova, J. Tilak Ratnanather, Qian-Li Xue, Susan M. Resnick, Yuri Agrawal

## Abstract

Emerging evidence suggests a relationship between impairments of the vestibular (inner ear balance) system and alterations in the function and the structure of the central nervous system in older adults. However, it is unclear whether age-related vestibular loss is associated with volume loss in brain regions known to receive vestibular input. To address this gap, we investigated the association between vestibular function and the volumes of four structures that process vestibular information (the hippocampus, entorhinal cortex, thalamus, and basal ganglia) in a longitudinal study of 97 healthy, older participants from the Baltimore Longitudinal Study of Aging. Vestibular testing included cervical vestibular-evoked myogenic potentials (cVEMP) to measure saccular function, ocular VEMP (oVEMP) to measure utricular function, and video head-impulse tests to measure the horizontal semi-circular canal vestibulo-ocular reflex (VOR). Participants in the sample had vestibular and brain MRI data for a total of 1 (18.6%), 2 (49.5%) and 3 (32.0%) visits. Linear mixed-effects regression was used to model regional volume over time as a function of vestibular physiological function, correcting for age, sex, intracranial volume, and inter-subject random variation in the baseline levels of and rates of change of volume over time. We found that poorer saccular function, characterized by lower cVEMP amplitude, is associated with reduced bilateral volumes of the basal ganglia and thalamus at each time point, demonstrated by a 0.0714 cm^3^ ± 0.0344 (unadjusted p=0.038; 95% CI: 0.00397-0.139) lower bilateral-mean volume of the basal ganglia and a 0.0440 cm^3^ ± 0.0221 (unadjusted p=0.046; 95% CI: 0.000727-0.0873) lower bilateral-mean volume of the thalamus for each 1-unit lower cVEMP amplitude. We also found a relationship between a lower mean VOR gain and lower left hippocampal volume (*β*=0.121, unadjusted p=0.018, 95% CI: 0.0212-0.222). There were no significant associations between volume and oVEMP. These findings provide insight into the specific brain structures that undergo atrophy in the context of age-related loss of peripheral vestibular function.

**Comprehensive Summary:** Humans rely on their vestibular, or inner ear balance, system to manage everyday life. In addition to sensing head motion and head position with respect to gravity, the vestibular system helps to maintain balance and gaze stability. Furthermore, evidence is mounting that vestibular function is linked to structural changes in the central nervous system (CNS). Yet, the exact processes by which vestibular function alters brain structural integrity is unclear. One possible mechanism is that progressive vestibular deafferentation results in neurodegeneration of structures that receive vestibular input. In support of this putative mechanism, recent studies report the association of vestibular impairment with volume loss of brain areas that receive vestibular information, specifically the hippocampus and entorhinal cortex, in older adults. This present work investigates the extent over time to which age-related vestibular loss contributes to volume reduction of four brain regions that receive vestibular input: the hippocampus, entorhinal cortex, thalamus, and basal ganglia. Using data from a cohort of healthy, older adults between 2013 and 2017 from the Baltimore Longitudinal Study of Aging, we assessed regional brain volume as a function of vestibular function, while accounting for common confounds of brain volume change (e.g., age, sex, head size). We found that poor vestibular function is associated with significantly reduced volumes of the thalamus, basal ganglia, and left hippocampus. Notably, this study is one of the first to demonstrate relationships between age-related vestibular loss and gray matter loss in brain regions that receive vestibular input. Further research is needed to understand in greater detail the observed link between vestibular function and CNS structure. Which brain areas are impacted by age-related vestibular loss? How and in what sequence are they impacted? As the world’s aging population—and the prevalence of age-related vestibular impairment—increases, answering questions like these becomes increasingly important. One day, these answers will provide targets for preemptive interventions, like physical pre-habilitation, to stave off adverse changes in brain structure before they occur and progress towards clinical significance.

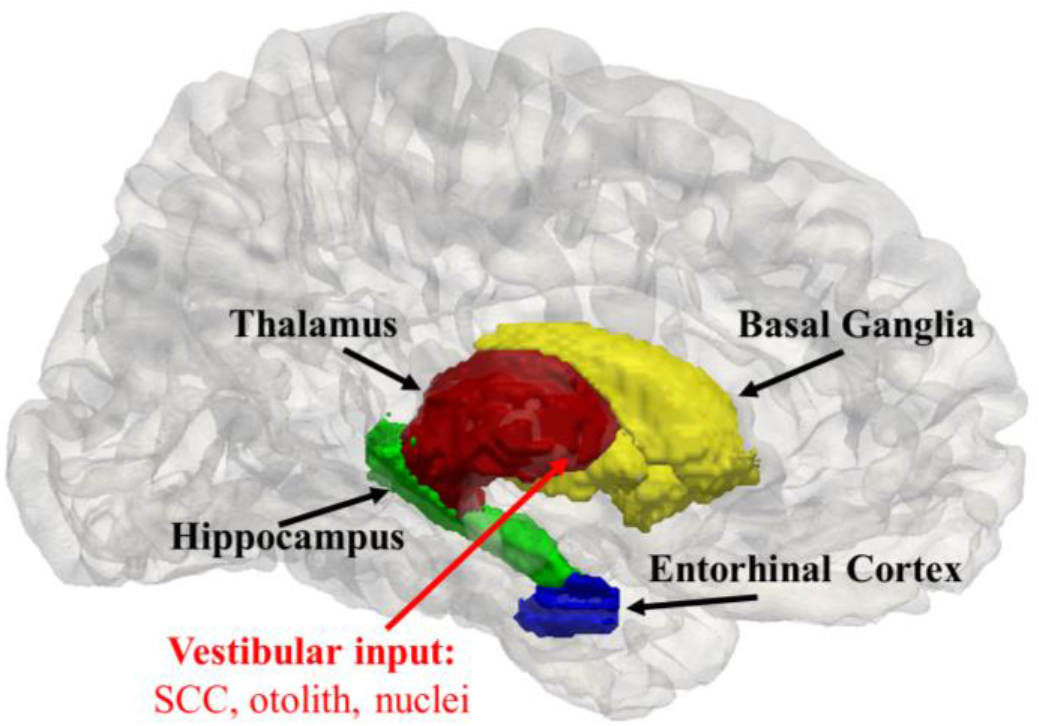

## 1. Introduction

The vestibular (inner ear balance) system consists of semi-circular canals which detect rotational movements of the head, and otolith organs, which detect linear head movements and the orientation of the head with respect to gravity [19]. The vestibular sensory system is known to play a critical role in maintaining stable balance and gaze control. Additionally, a growing body of literature indicates a pre-eminent contribution of the vestibular system to visuospatial cognitive ability [1,19-22]. Vestibular function declines with normal aging [23-25], and vestibular dysfunction is associated with cognitive declines in older adults [1,17,18], including in spatial cognitive function [26,27].

Functional neuroimaging and vestibular stimulation studies have demonstrated that vestibular information is transmitted to multiple neural regions including, the hippocampus, thalamus, basal ganglia, entorhinal cortex, cerebellum, insula, parietal regions, and somatosensory regions [3-5,28-37]. In fact, several clinical studies have shown that lack of vestibular input leads to functional and structural changes in the brain, either directly or indirectly. A pioneering study of subjects with bilateral vestibulopathy (BVP) by Brandt et al. [32] demonstrated that lack of vestibular input leads to bilateral hippocampal atrophy (16.91% reduction relative to controls) that correlated with impaired spatial memory and navigation assessed by the virtual Morris water navigation task (vMWT). Hufner et al. [47] reported reductions in gray matter volume in the cerebellum due to the schwannoma removal, supramarginal gyrus on the same side as the lesion, postcentral and superior temporal gyri, and in motion-sensitive visual area MT/V5 in patients with complete unilateral vestibular deafferentation (UVD) compared to healthy controls. Helmchen et al. [50] demonstrated increases in gray matter volume in the insula, retroinsular vestibular cortex, and superior temporal gyrus correlated with improving clinical vestibular assessments in patients recovering from vestibular neuritis. zu Eulenberg et al. [49] showed that patients with an acute unilateral vestibular deficit stemming from vestibular neuritis recovery exhibited gray matter volume reduction in the left posterior hippocampus, the right superior temporal gyrus, and the right superior frontal gyrus regardless of the side of vestibular impairment. Göttlich et al. [45] used voxel-based morphometry (VBM) to find reductions in gray-matter volume in the CA3 region of the hippocampus bilaterally with increasing vestibulopathy-related disability in patients with incomplete BVP compared to healthy age- and gender-matched controls. Kremmyda et al. [48] found that vestibular loss due to incomplete BVP leads to gray matter volume reductions bilaterally in the middle hippocampus and posterior parahippocampus using VBM, to deficits in objective and subjective evaluations of spatial memory and navigation performance using the vMWT and questionnaires, and to increased spatial anxiety. In a cross-sectional VBM study of patients with persistent postural perceptual dizziness, Wurthmann et al. [46] demonstrated reductions in gray matter volumes in the left superior temporal gyrus, left motion-sensitive visual area MT/V5 and bilateral middle temporal gyrus, bilaterally in the cerebellum, left-sided posterior hippocampus, right precentral gyrus, left anterior cingulate cortex, left side of left caudate nucleus, and left dorsolateral prefrontal cortex.

However, little is known about which structures are affected by subclinical, age-related vestibular loss. Furthermore, little is known about the specific vestibular end-organs (i.e. semicircular canal and/or otolith organs) that contribute to those changes. Bridging these knowledge gaps, novel cross-sectional studies in healthy, older individuals found significant associations between age-related vestibular loss and gross gray matter loss of the hippocampus [2,41] and the entorhinal cortex [41], as well as shape changes in the hippocampus, amygdala, caudate nucleus, putamen, thalamus, entorhinal cortex, and the entorhinal-transentorhinal cortical complex [41].

In this study, we examined the relationship between age-related vestibular function and regional volume of the hippocampus, entorhinal cortex, thalamus, and basal ganglia, four structures known to process vestibular information. Linear mixed-effects regression was used to evaluate the association between saccular, utricular, and horizontal semi-circular canal vestibulo-ocular reflex functions and MRI-based volumes of 97 of healthy, older participants aged 60 years and older from the Baltimore Longitudinal Study of Aging over a five-year follow-up period. To our knowledge, this study is one of the first to investigate the associations over time between age-related vestibular function and volumes of brain regions that receive vestibular input.

## 2. Materials and Methods

### 2.1. Data

Data on 97 older adult participants (aged ≥ 60 years) who had at least one vestibular physiological test and structural MRI scan on the same visit between 2013 and 2017 were selected from the Baltimore Longitudinal Study of Aging (BLSA) [6,44]. All participants gave written informed consent, and none had any history of psychiatric disorders or were diagnosed with a vestibular, ophthalmological, or neurodegenerative disease. Hearing loss was measured and was included as a confounding variable in the supplemental analysis.

### 2.2. Vestibular physiologic testing

Vestibular function testing was based on the cervical vestibular-evoked myogenic potential (cVEMP), ocular VEMP (oVEMP), and video head-impulse testing (vHIT), which measure saccular function, utricular function, and the horizontal semicircular canal vestibular-ocular reflex (VOR), respectively. cVEMP and oVEMP were recorded using a commercial electromyographic system (software version 14.1, Carefusion Synergy, Dublin, OH) [7,8]. Electromyograms (EMG) were recorded by disposable, pre-gelled Ag/AgCl electrodes placed with 40-inch safety lead wires from GN Otometrics (Schaumburg, IL). cVEMP and oVEMP signals were amplified ×2500 and band-pass filtered for the 20-2000 Hz and 3-500 Hz frequency intervals, respectively.

#### 2.2.1. Cervical vestibular-evoked myogenic potential (cVEMP)

cVEMPs are short latency EMGs of the relaxation response of the sternocleidomastoid muscles induced by sound stimuli in the ear or by head-tapping vibrations. They measure saccular (and inferior vestibular nerve) function. The saccule is a vestibular end-organ that transduces linear acceleration, detects the orientation of the head with respect to gravity, and plays a role in spatial cognition [8]. Testing followed an established protocol [7,8,9,10]. Participants sat on a chair inclined to 30° above the horizontal plane, and qualified examiners placed EMG electrodes on the sternocleidomastoid and sternoclavicular junction bilaterally and a ground electrode on the manubrium sterni. Sound stimuli of 500 Hz and 125 dB were administered in bursts of 100 stimuli monoaurally through headphones. Recorded cVEMP amplitudes were corrected for spontaneous background EMG activity collected 10 ms prior to the onset of the sound stimulus. An absent response was defined according to previously published amplitude and latency thresholds [7,8]. In case of an absent recording, the response assessment was repeated for confirmation. For present responses, the higher cVEMP from the left and right sides was used in this analysis.

#### 2.2.2. Ocular vestibular-evoked myogenic potential (oVEMP)

oVEMPs are short latency EMGs of the excitation response of the inferior oblique muscles of the eye elicited by vibration stimulation of the skull. They measure utricular (and superior vestibular nerve) function [8]. The utricle is the otolith organ that transduces linear acceleration and detects the orientation of the head with respect to gravity. Testing followed an established protocol [7,8,9,10]. Participants sat on a chair inclined to 30°, and qualified examiners placed a noninverting electrode ∼3 mm below the eye centered below the pupil, an inverting electrode 2 cm below the noninverting electrode, and a ground electrode on the manubrium sterni. To ensure that the signals recorded from both eyes are symmetric before stimulation, participants were asked to perform multiple 20° vertical saccades. New electrodes were applied in place of the old ones if the signal difference exhibited >25% asymmetry. Participants were asked to maintain an upgaze of 20° during oVEMP testing and recording. Head taps were applied to the midline of the face at the hairline and ∼30% of the distance between the inion and nasion using a reflex hammer (Aesculap model ACO12C, Center Valley, PA). An absent response was defined according to previously published amplitude and latency thresholds [7,8]. In case of an absent recording, the response assessment was repeated for confirmation. For present responses, the higher oVEMP from the left and right sides was used in this analysis.

#### 2.2.3. Video head impulse test (vHIT)

The horizontal vestibular-ocular reflex (VOR) was assessed using the vHIT [10,14,15]. To determine VOR gain, the vHIT was performed using the EyeSeeCam system (Interacoustics, Eden Prarie, MN) in the same plane as the right and left horizontal semicircular canals [11,15]. The participant’s head was tilted downward 30° below the horizontal plane to correctly position the horizontal canals in the plane of stimulation. Participants were asked to maintain their gaze on a wall target ∼1.5 m away. A qualified examiner rotated the participant’s head 5-10° quickly (∼150-250° per second) parallel to the ground toward the right and left at least 10 times in both directions, chosen randomly for unpredictability. The EyeSeeCam system quantified eye and head velocity, and a corresponding VOR gain was calculated as the unitless ratio of the eye velocity to the head velocity. A normal eye and head velocity should be equal, yielding a VOR gain equal to 1.0. A VOR gain <0.8 accompanied by refixation saccades suggests peripheral vestibular hypofunction [12,14]. The mean VOR gain from the left and right sides was used in this study.

### 2.3. Structural MRI acquisition and processing

T1-weighted volumetric MRI scans were acquired in the sagittal plane using a 3T Philips Achieva scanner at the National Institute on Aging Clinical Research Unit. Sequences included a T1 volumetric scan magnetization prepared rapid acquisition with gradient echo (MPRAGE; repetition time (TR)=6.5 ms, echo time (TE)=3.1 ms, flip angle=8°, image matrix=256×256, 170 slices, voxel size=1.0×1.0×1.2 mm slice thickness, FOV=256×240 mm, sagittal acquisition). A semi-automated quality control protocol was employed to automatically identify and manually exclude scans which have segmentation errors defined as outliers of each region-specific sample distribution. Anatomical labels were obtained using Multi-atlas region Segmentation using Ensembles of deformable registration algorithms and parameters [39]. Global and regional brain volumes were calculated from the binary segmentations output by MUSE by counting the total number of voxels in the binary label image and multiplying that voxel count by the spatial size of a voxel in *cm*^3^. We corrected for intracranial volume (ICV) individually estimated at age 70 using the residual volume approach described by Jack et al. [40]: for each region and each scan, residual volumes were calculated as the difference, in cm^3^, of the measured regional volume from the expected regional volume, given the ICV for the individual. The basal ganglia volume was defined to be the sum of the volumes of the lenticular nucleus, globus pallidus, and the caudate nucleus. Figure 1 provides a visualization of the relative locations of the thalamus, hippocampus, basal ganglia, and entorhinal cortex in a three-dimensional hemi-brain.

**Figure 1.**
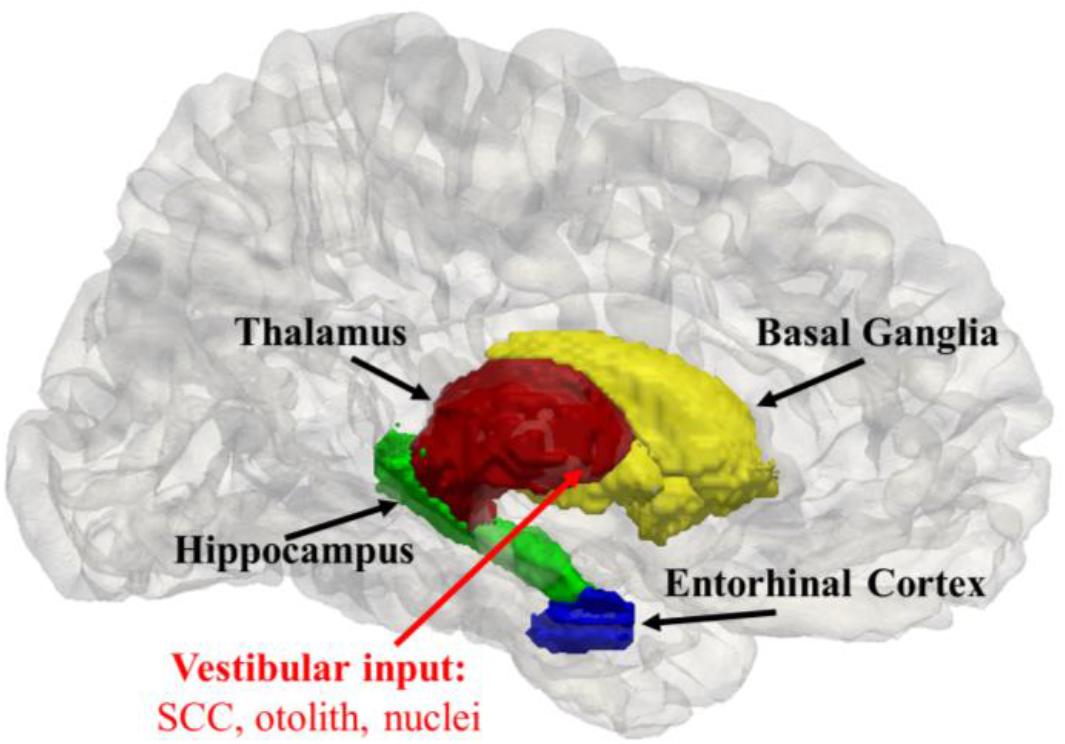
Simplified 3D illustration of three sub-cortical and one cortical regions of the vestibular network in the left hemisphere of the JHU-MNI-SS brain [42]. The medial view of the left side of the pial surface of the JHU-MNI-SS template [42] generated by FreeSurfer and mapped to native space [43] is shown. The three subcortical (thalamus, hippocampus, basal ganglia, comprised of the putamen, caudate nucleus, and globus pallidus) and one cortical (entorhinal cortex) structures were obtained from the JHU-MNI-SS labels and triangulated. Vestibular information from the semi-circular canals (SCC), otoliths, and nuclei is relayed through the thalamus to the hippocampus, entorhinal cortex, and basal ganglia. The red arrow points toward the ventral lateral nucleus, a putative subfield of the thalamus that receives vestibular input [41]. CAWorks (www.cis.jhu.edu/software/caworks) was used for visualization.

### 2.4. Mixed-effects modeling

Linear mixed-effects regression was used to model the evolution of regional brain volume over time while allowing a time-independent cross-sectional effect of vestibular function on regional brain volume. The model included a by-subject random intercept and a random slope on age to account for inter-subject heterogeneity in the baseline level of and rate of change in regional brain volume over time.

The null hypothesis in Eq. (1) predicts regional volume *vol*_*i,j*_, for participant *i, i* = 1, …, *N* for observation *j, j* = 1, …, *n*_*i*_. The alternate hypotheses predict volume using mean VOR gain termed *VOR*_*i,j*_ in Eq. (2), *oVEMP*_*i,j*_ in Eq. (3), and *cVEMP*_*i,j*_ in Eq. (4) as continuous independent variables,

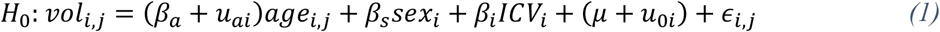

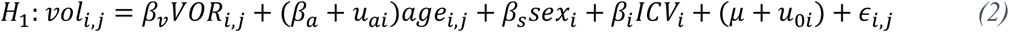

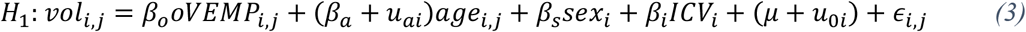

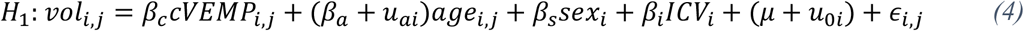

where *age*_*i,j*_ of subject *i* at visit *j* is centered at age 70, *sex*_*i*_ is a binary indicator variable for the sex of subject *i*, and *ICV*_*i*_ denotes baseline intracranial volume, comprised of bilateral cerebral volumes, cerebellum, brainstem, and cerebrospinal fluid, individually estimated at 70 years of age. Age, sex, and intracranial volume were treated as fixed effects. We assumed linear dependence of regional volume with respect to age. We assumed *u*_*ai*_, *u*_0*i*_ are zero-mean Gaussian distributed with unknown variances and covariances and respectively represent the intra-subject random effect, which captures correlations between measurements within subject *i* over time, and the between-subject heterogeneity in terms of individual deviation in regional volume from its sample mean at age 70. The covariance structure of random effects was taken to be unstructured. We assumed measurement noise *ϵ*_*i,j*_ is independently and identically distributed zero-mean Gaussian with unknown common variance. The unknown fixed-effects { *β*_*v*_, *β*_*o*_, *β*_*c*_, *β*_*a*_, *β*_*s*_, *β*_*i*_, *μ* } and random-effects covariance matrix parameters were estimated via maximum likelihood. All effects were considered significant at the p<0.05 level. A Benjamini-Hotchberg procedure was used to control the false discovery rate (FDR) of the comparisons made in this study [38]. FDR q-values indicate the expected proportion of rejected null hypotheses that are false. We considered an FDR threshold of 0.05 and also 0.10, given that these were initial analyses testing specific hypotheses based on prior work. These statistics were performed using the *xtmixed* function in Stata 15 (College Station, Texas).

## 3. Results

### 3.1. Baseline and longitudinal characteristics

Table 1 shows the baseline characteristics for the study sample from the Baltimore Longitudinal Study of Aging. Eighteen participants had one observation with both vestibular and MRI data available, forty-eight participants had two observations, and thirty-one participants had three observations. In this sample, there was only a single case of participant dropout, due to death, after two visits. At baseline, this participant had low cVEMP, missing oVEMP and VOR, and an average hippocampal volume.

**Table 1.** Baseline characteristics of the study sample (n = 97). Regional volumes are given in cm^3^. Observation Count denotes the number of participants with at least one visit where both vestibular and MRI data were available. Key: SD: standard deviation.

| Characteristic | Overall |
| --- | --- |
| N | 97 |
| Age (years), mean (SD) | 76 (8.39) |
| Sex (% female) | 58.76 |
| Region, mean (SD) |  |
| Intracranial Volume | 1391.39 (147.32) |
| Hippocampus | 7.23 (0.81) |
| Thalamus | 14.2 (1.41) |
| Basal Ganglia | 17.9 (1.99) |
| **Entorhinal Cortex** | 4.29 (0.605) |
| **Observation Count, n (%)** |  |
| **1** | 18 (18.6) |
| **2** | 48 (49.5) |
| **3** | 31 (32.0) |

### 3.2. Association between vestibular function and regional brain volumes based in mixed-effects models

Table 2 reports the association between vestibular function and regional brain volume. We found significant relationships in which each 1-unit lower cVEMP amplitude was associated with a 0.0714 cm^3^± 0.0344 (unadjusted p=0.038, 95% CI: 0.00397-0.139) lower bilateral-mean volume of the basal ganglia and a 0.0440 cm^3^ ± 0.0221 (unadjusted p=0.046, 95% CI: 0.000727-0.0873) lower bilateral-mean volume of the thalamus. This means every one-unit decrease in cVEMP amplitude is associated with an average reduction of ∼0.4% in the basal ganglia and ∼0.31% in the thalamus. We examined the left and right hemispheres separately and found significant relationships for the left thalamus (*β*=0.0232, unadjusted p=0.043, 95% CI: 0.000684-0.0457) and for both the left (*β*=0.0376, unadjusted p=0.048, 95% CI: 0.000385-0.0747) and right (*β*=0.0359, unadjusted p=0.045, 95% CI: 0.000727-0.071) basal ganglia. The effect was borderline for the right thalamus (*β*=0.0231, unadjusted p=0.065, 95% CI: -0.00148-0.0477). We also found a significant relationship between a lower mean VOR gain and lower left hippocampal volume (*β*=0.121, unadjusted p=0.018, 95% CI: 0.0212-0.222). In other words, every one-unit decrease in mean VOR gain is associated with an average reduction of 0.121 cm^3^, or ∼1.68%, in the left hippocampus. We found no significant relationships between cVEMP and volume of the hippocampus or the entorhinal cortex, no significant associations between VOR and brain volume aside from the left hippocampus, and no significant associations between oVEMP and regional volume. After controlling the FDR at the threshold of 0.10, lower cVEMP amplitude was associated with lower bilateral-mean volume of the thalamus (q = 0.092 FDR corrected) and with lower bilateral-mean volume of the basal ganglia (q = 0.092 FDR corrected). In additional analyses, we added hearing – represented by the for 4-frequency pure tone average from the better ear – to the regression models. The addition of hearing to the models reduced the sample size from 97 to 84 participants and resulted in the significant associations previously observed becoming marginally significant, although with similar effect sizes (See Supplementary Table S1).

**Table 2.**
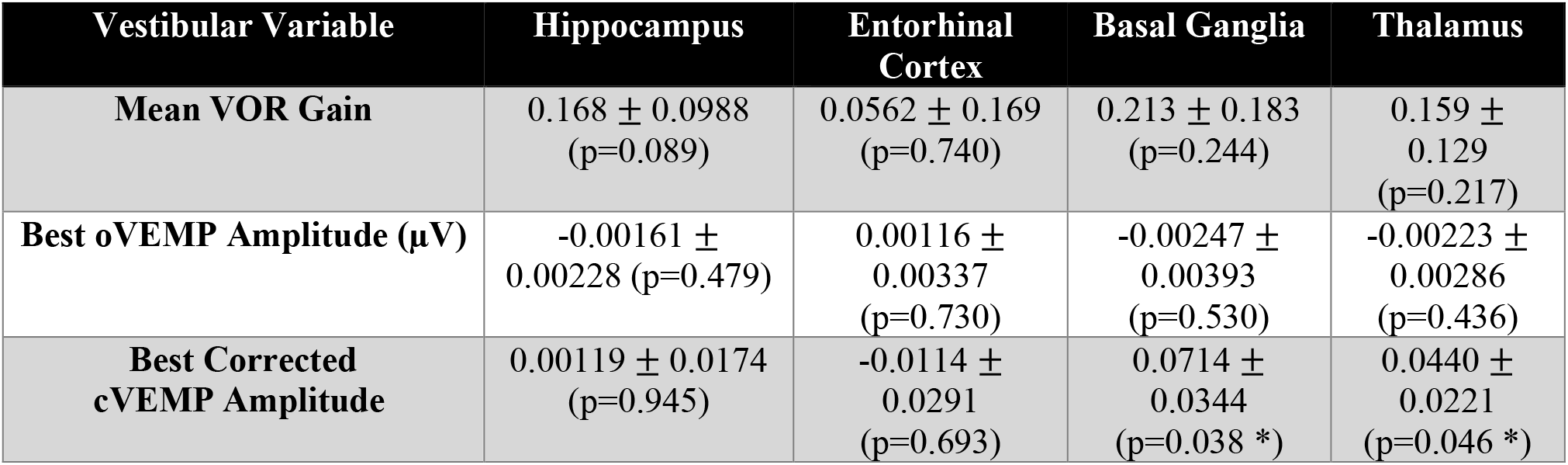
Vestibular predictors of regional volume (cm^3^) under the alternate hypothesis with mean ± standard error (unadjusted p-value). Key: *: p < 0.05.

The “aging” effect was significant for all models (q << 0.0001 FDR corrected), except for the left (*β*=-0.0169, unadjusted p=0.104), right (*β*=-0.0167, unadjusted p=0.064), and bilateral-mean (*β*=-0.0348, unadjusted p=0.064) basal ganglia cVEMP models, which showed strong trends of volume reductions. For the thalamus cVEMP model, every one-year increase in age was associated with a 0.0778 cm^3^ ± 0.00834 (q << 0.0001 FDR corrected, 95% CI: (−0.0941, -0.0614)), or -0.55% per year, reduction in bilateral-mean volume and a 0.0392 cm^3^ ± 0.0041 (q << 0.0001 FDR corrected, 95% CI: (−0.0472, -0.0311)) reduction in left-side volume (−0.27%/year). For the left hippocampus VOR model, every one-year increase in age was associated with a 0.0239 cm^3^ ± 0.00271 (q << 0.0001 FDR corrected, 95% CI: (−0.0292, -0.0186)) reduction in volume (−0.33%/year). Sex predicted entorhinal cortex volume over time in each model (q << 0.0001 FDR corrected) and right thalamus volumes in the VOR model (*β*=0.248, unadjusted p=0.045, 95% CI: (0.00539, 0.49)). Despite failing to reject the null hypothesis, sex showed strong trends for the bilateral-mean (*β*=0.485, unadjusted p=0.052) and left (*β*=0.23, unadjusted p=0.073) thalamus VOR models, and was not a predictor of volume trajectories for the hippocampus, basal ganglia, or the thalamus cVEMP or oVEMP models.

Because age is included as a covariate, the volume growth trajectory related to vestibular function represents effects above and beyond those attributable to normal age-related gray matter volume loss. This means, in addition to the age-related volume reduction rate of 0.19% per year in the basal ganglia and 0.55% per year in the thalamus, every one-unit decrease in cVEMP amplitude is associated with an average reduction of ∼0.4% in the basal ganglia and ∼0.31% in the thalamus. In other words, the cVEMP effects are 2.05 times larger in magnitude and 0.57 times smaller in magnitude than the age-related volume reductions in the basal ganglia and thalamus, respectively. The VOR effects are 5.08 times larger in magnitude than the age-related volume reduction rate in the left hippocampus.

## 4. Discussion

These findings provide some of the first evidence for associations between lower cVEMP amplitude, indicative of poorer saccular function, and significantly reduced volumes of the thalamus and basal ganglia and lower VOR gain (poorer canal function) and reduced left hippocampal volumes over time in healthy, older adults aged 60 years and older. These results are compatible with clinical studies in patients with vestibular loss which demonstrated that loss of peripheral vestibular inputs is associated with gray matter reductions in the hippocampus [32,45,46,48,49] and basal ganglia [46]. cVEMP measures the physiological function of the saccule, the vestibular end-organ that detects linear acceleration and gravitational cues and preserves spatial orientation [8,19]. VOR gain measures the physiological function of the horizontal semi-circular canal, the vestibular end-organ that detects angular acceleration in the transverse plane [19]. Prior work has shown that abnormal VOR gain [26] and lower cVEMP amplitude are significantly associated with poorer spatial cognitive performance in healthy, older adults [19,26,27]. Importantly, this work expands the understanding of the role of the peripheral vestibular system in anatomical alterations.

We note that the effect sizes of the relationship between vestibular measures and changes in gray matter volumes over time are small. These small though significant effect sizes may reflect the indirect links between the peripheral vestibular system and the thalamus and basal ganglia. Vestibular inputs are known to traverse through brain structures such as the cerebellum, brainstem, and hypothalamus, where additional afferent inputs are integrated (e.g. visual, auditory, autonomic), thereby modulating and attenuating the direct impact of vestibular loss on central structural changes [53,54]. Whether these changes in gray matter volumes represent loss of neurons remains unclear in the absence of additional physiological or tissue studies. As such, the extent to which these effect sizes reflect underlying neuronal loss is unclear. Notably, the aging effects described are consistent with the age-related cerebral volume reduction rates of ∼0.5% per year found in previous longitudinal studies in normal older controls [51,52].

A previous cross-sectional study in healthy, older adults demonstrated that saccular sensory loss, defined as lower cVEMP amplitude, was associated with significant gross volume loss of the hippocampus [2]. We did not observe the same relationship between cVEMP amplitude and hippocampal volume in this longitudinal analysis. A possible explanation for this discrepancy between the cross-sectional and longitudinal findings is that the hippocampal volume reduction may have largely occurred prior to the start of this longitudinal period, such that the relationship was significant in the baseline cross-sectional analysis but not in the longitudinal analysis. In support of this explanation, one-way ANCOVA controlling for age, sex, and intracranial volume suggests the baseline bilateral-mean hippocampal volume in participants with impaired cVEMP at baseline is 0.34 cm^3^ smaller compared to those with unimpaired cVEMP at baseline (p=0.014, 95% CI: (−0.608, -0.0717)). Another possible explanation for the discrepancy between our cross-sectional and longitudinal findings is a systematic difference between the two cohorts, due to non-random, biased loss to longitudinal follow-up. However, this is unlikely, given that there was only a single participant who dropped out of this study sample after two visits due to death. Additionally, there was no difference in the proportion of participants with normal vs. low cVEMP amplitude (≤25^th^ percentile) at baseline who had intermittently missing follow-up observations (and in all cases all available data were used in analyses).

We note limitations of this study. The relationship between vestibular function and brainstem and cerebellar structures were not studied, as these structures have been more challenging to parcellate due to their complex anatomy, and robust measures to analyze the brainstem and cerebellum in all BLSA participants are currently being developed. Additionally, we also did not investigate the potential mediation or modification of the effects of age-related vestibular loss by intervening brain structures like the cerebellum, the brainstem, or the hypothalamus. Moreover, the generalizability of our results is limited by the age range studied and the tendency of the BLSA participants to have higher levels of education and socioeconomic status than the broader adult population. Whether these findings can also be detected in younger adults with reduced vestibular function is unclear.

Future work will be needed to further clarify the relationships between vestibular function and the structure of regions of the limbic system, temporo-parietal junction, and frontal cortex – all of which receive vestibular input – and their temporal sequence of effects relative to each other. Additionally, subsequent studies will need to investigate the direction of causal influence between vestibular loss and regional size and shape changes, while correcting for potential confounding factors such as hearing or vision loss. Structural equation modeling with longitudinal data in which vestibular measurements precede structural measurements can help tease out the direct or indirect relationships between vestibular function and brain structures. Cortical thickness of cortical structures, such as the entorhinal cortex, insula, and prefrontal cortex, can provide a sensitive measure of cortical integrity complementary to shape changes. Changepoint analysis of longitudinal data can identify non-linearities in the trajectories of structural change. By temporally ordering the set of changepoints for each structure measure, the sequence of changes in the vestibular network can be revealed. Future studies will be needed to test whether there are modulating effects on downstream structures via intervening brain structures such as the brainstem, the hypothalamus, and the cerebellum.

## 5. Conclusions

This study examined the association between reduced vestibular function and regional brain volume over time in aging adults. To our knowledge, this study is one of the first to demonstrate significant relationships between vestibular loss—saccular and semicircular canal sensory loss in particular—and gray matter volume loss of the thalamus, basal ganglia, and left hippocampus, three vestibular subcortical structures that receive peripheral vestibular input. Future work will need to determine the timing and sequence of the relationships between vestibular function and neuromorphological alterations.

## Data Availability

This study used data that is publicly available through the Investigator's portal of the Baltimore Longitudinal Study of Aging.

## Acknowledgements

This work was supported in part by the National Institute on Aging [grant number R01 AG057667], National Institute on Deafness and Other Communication Disorders [grant number R03 DC015583], and National Institute of Health [grant number P41 EB015909] and by the Intramural Research Program, National Institute on Aging, National Institutes of health.

