## Supplemental Table S1 for "Linking vestibular function and sub-cortical grey matter volume changes in a longitudinal study of aging adults"

**Supplemental Information**

**Table S1.** Vestibular and hearing predictors of regional volume ($cm^3$) under the alternate hypothesis additionally controlling for the four-frequency pure tone average (PTA) (n=84) with mean ± standard error (uncorrected p-value). The speech-frequency PTA of air-conduction thresholds at 0.5, 1, 2, and 4 kHz from the better ear was used. Key: PTA: Pure Tone Average, *: p < 0.05, **: p < 0.01.

| Vestibular Variable | Outcome | Effect of Vestibular Variable on Outcome | Effect of PTA on Outcome |
|---|---|---|---|
| **Best corrected cVEMP amplitude** | | | |
| | Hippocampus | -0.0185 ± 0.0181 (p=0.311) | -0.00844 ± 0.00428 (p=0.0517) |
| | Left | -0.00210 ± 0.00916 (p=0.82) | -0.00372 ± 0.00214 (p=0.0856) |
| | Right | -0.0162 ± 0.0112 (p=0.154) | -0.00456 ± 0.00253 (p=0.0740) |
| | Thalamus | 0.0380 ± 0.0244 (p=0.125) | 0.00371 ± 0.00600 (p=0.538) |
| | Left | 0.0191 ± 0.0127 (p=0.138) | 0.000769 ± 0.00312 (p=0.806) |
| | Right | 0.0192 ± 0.0134 (p=0.159) | 0.00253 ± 0.00320 (p=0.431) |
| | Basal Ganglia | 0.0653 ± 0.0335 (p=0.0596) | 0.0108 ± 0.00889 (p=0.230) |
| | Left | 0.0292 ± 0.0173 (p=0.106) | 0.00563 ± 0.00456 (p=0.224) |
| | Right | 0.0332 ± 0.0185 (p=0.0798) | 0.00503 ± 0.00479 (p=0.297) |
| | Entorhinal Cortex | -0.0306 ± 0.0322 (p=0.344) | 0.00739 ± 0.00503 (p=0.144) |
| | Left | -0.0243 ± 0.0179 (p=0.179) | 0.00404 ± 0.00300 (p=0.181) |
| | Right | -0.00577 ± 0.0191 (p=0.763) | 0.00389 ± 0.00265 (p=0.145) |
| **Best oVEMP Amplitude (μV)** | | | |
| | Hippocampus | -0.00146 ± 0.00216 (p=0.501) | -0.0057 ± 0.00393 (p=0.149) |
| | Left | -0.000629 ± 0.00118 (p=0.597) | -0.00392 ± 0.00205 (p=0.058) |
| | Right | -0.000887 ± 0.00125 (p=0.479) | -0.00159 ± 0.00224 (p=0.479) |
| | Thalamus | -0.0026 ± 0.00295 (p=0.383) | 0.00878 ± 0.00551 (p=0.115) |
| | Left | -0.000943 ± 0.00159 (p=0.555) | 0.00374 ± 0.00293 (p=0.206) |
| | Right | -0.00148 ± 0.00161 (p=0.363) | 0.00478 ± 0.00294 (p=0.108) |

| | | | |
|---|---|---|---|
| | Basal Ganglia | -0.00213 ± 0.00444 (p=0.634) | 0.0154 ± 0.00891 (p=0.0879) |
| | Left | -0.0015 ± 0.00198 (p=0.453) | 0.00708 ± 0.0041 (p=0.0889) |
| | Right | -9.39e-05 ± 0.00267 (p=0.972) | 0.00767 ± 0.00511 (p=0.137) |
| | Entorhinal Cortex | 0.000749 ± 0.00345 (p=0.829) | 0.00893 ± 0.00455 (p=0.0519) |
| | Left | 0.00176 ± 0.00187 (p=0.351) | 0.00498 ± 0.00273 (p=0.0709) |
| | Right | -0.000533 ± 0.00195 (p=0.785) | 0.00481 ± 0.00227 (p=0.0359 *) |
| **Mean VOR Gain** | | | |
| | Hippocampus | 0.217 ± 0.096 (p=0.0273 *) | -0.00574 ± 0.00359 (p=0.113) |
| | Left | 0.138 ± 0.0494 (p=0.00697 **) | -0.00308 ± 0.00181 (p=0.0914) |
| | Right | 0.076 ± 0.0554 (p=0.175) | -0.00257 ± 0.00205 (p=0.213) |
| | Thalamus | 0.181 ± 0.123 (p=0.146) | 0.00431 ± 0.00483 (p=0.375) |
| | Left | 0.121 ± 0.068 (p=0.0797) | 0.00316 ± 0.00263 (p=0.233) |
| | Right | 0.057 ± 0.0676 (p=0.402) | 0.00114 ± 0.00259 (p=0.662) |
| | Basal Ganglia | 0.189 ± 0.19 (p=0.325) | 0.0117 ± 0.00753 (p=0.122) |
| | Left | 0.129 ± 0.0945 (p=0.178) | 0.00737 ± 0.00379 (p=0.0552) |
| | Right | 0.0564 ± 0.113 (p=0.619) | 0.00455 ± 0.00421 (p=0.283) |
| | Entorhinal Cortex | -0.0259 ± 0.177 (p=0.884) | 0.00691 ± 0.00431 (p=0.111) |
| | Left | 0.0316 ± 0.0994 (p=0.752) | 0.00289 ± 0.00264 (p=0.276) |
| | Right | -0.053 ± 0.094 (p=0.574) | 0.00423 ± 0.00209 (p=0.0459 *) |
